# Micro-costing for national-scale azithromycin mass drug administration to improve child survival in Niger

**DOI:** 10.64898/2026.02.09.26345952

**Authors:** Brittany Peterson, William Nguyen, Laminou Maliki Haroun, Farissatou Oumarou, Ahmed M. Arzika, Ramatou Maliki, Abdou Amza, Karamba Alio, Nasser Gallo, Bawa Aichatou, Ismael Issa Sara, Diallo Beidi, James G. Kahn, Stefano M. Bertozzi, Elodie Lebas, Benjamin F. Arnold, Thomas M. Lietman, Kieran S. O’Brien, Meagan C. Fitzpatrick, the AVENIR Study Group

## Abstract

As programs for azithromycin mass drug administration to reduce child mortality have begun in some parts of West Africa, it is imperative to understand their financial costs. We combined a micro-costing framework and observations from an implementation-focused sub-study within the AVENIR trials in 80 communities in the Dosso region of Niger to estimate the national health sector costs of a scaled-up programmatic approach for azithromycin biannual distribution to children aged 1-59 months of age living in nonurban areas, using the door-to-door modality. Our outcomes of interest were the annual budget at the regional and national levels for Niger and the cost per dose delivered. We found that the annual national budget required for azithromycin mass drug administration (MDA) achieving 90% average coverage would be $12.5M (95% Uncertainty Interval (UI) $12.2M, $13.0M) translating to $1.59 (95% UI $1.40, $2.30) per dose delivered. Across regions, cost per dose would vary from $1.17 (95% UI $1.03, $1.69) to $3.61 (95% UI $3.20, $5.16), with higher cost per dose expected for more sparsely populated regions. Training costs represented a large fraction (16.4%) of total costs, and integration of training with that for existing health interventions may provide opportunities for efficiency.

## INTRODUCTION

Under-5 mortality (U5MR) in the West and Central Africa region remains much higher than United Nations Sustainable Development Goal 2030 targets.^1–3^ The region reports 95 deaths per 1000 live births in 2019, which is 19 times higher than the U5MR reported in high income settings.^1,2^ Azithromycin mass drug administration (AZ-MDA) is one effective approach to reduce child mortality. The *Macrolides Oraux pour Réduire les Décès avec un Oeil sur la Résistance* trial (MORDOR) found that biannual distribution of oral azithromycin to children 1-59 months old reduced mortality by 14% in Malawi, Niger, and Tanzania after 2 years, with longer-term follow-up confirming these findings.^4–6^ In 2020, the World Health Organization released conditional guidelines suggesting this intervention be considered in some high mortality settings in sub-Saharan Africa.^7^ Recently, the AVENIR (*Azithromycine pour la Vie des Enfants au Niger: Implementation et Recherche*) trial in Niger confirmed the intervention effectiveness observed during MORDOR, leading to plans for broad roll-out of AZ-MDA in Niger, Mali, Nigeria, and potentially elsewhere.^8,9^

MORDOR and follow-up studies were resource-intensive and relied on dedicated study teams for azithromycin distribution. However, programmatic implementation will ultimately occur within existing health systems and must be affordable. Our team conducted a cluster-randomized trial to evaluate programmatic approaches for a standalone AZ-MDA that relied on community health workers (CHWs), comparing door-to-door and fixed-point delivery across several indicators, including reach, costs, fidelity, and acceptability at multiple stakeholder levels.^10^ The study found that although the door-to-door delivery modality had slightly higher costs, it also resulted in a greater number of doses delivered and was therefore more efficient (i.e., had a lower cost per dose).

The World Health Organization has estimated the cost of mass drug administration (MDA) for neglected tropical diseases to be less than $0.50 USD per person per year^11^. However, one systematic review of 34 studies evaluating the cost of MDA against neglected tropical disease found broad variation in on-the-ground costs.^12^ Within these 34 studies, 90% of MDA programs that included cost effectiveness analyses used local volunteer time, which provides financial savings but may compromise program coverage or reach^12^. In Niger, the use of volunteers for MDA is not common practice as payment ensures quality of work and availability of CHWs so all CHWs involved in MDA are paid for their time. In a regression model, the review found that the financial cost more than doubles when using paid health workers^12–14^. Furthermore, economies of scale were important, with less populated areas incurring a higher cost per dose. For instance, an MDA targeting trachoma control in remote areas of South Sudan cost $1.50 USD per person treated.^15^ Given the focus on non-urban areas for azithromycin, the use of paid CHWs, and currency inflation since the date of the review, we anticipate that the commonly-cited estimate of $0.50 per dose might substantially underestimate the financial resources required for AZ-MDA in Niger. Confirming this heterogeneity, a recent review of costs for community health care worker programs found a range spanning from $0.02 to $482 per beneficiary.^16^

The objective of this analysis was to use data from AVENIR to consider the budgetary implications at the regional and national levels as well as the cost per dose of azithromycin mass distribution to improve child survival. As effectiveness results become available from MDA demonstration projects in the region, this service delivery efficiency information will be crucial for policymakers contemplating program design and scale.

## METHODS

The AVENIR project included several studies on azithromycin mass distribution to improve child survival in Niger.^8,9^ The AVENIR Delivery I trial compared door-to-door and fixed point delivery. Because the door-to-door modality reached more children at a lower cost per child, it is the focus of our current study. Details regarding Delivery I trial design, ethics and oversight, interventions, adverse events specification, and data collection are reported in a linked paper.^10^ Briefly, trial approval was obtained from the Institutional Review Boards at the Niger Ministry of Health (Comité Nationale Éthique pour la Recherche en Santé) and the University of California, San Francisco. Trial recruitment began in November 2020. For the community level, local leaders provided verbal consent; at the individual level, caregivers provided written consent for children 30-42 days old and verbal consent for older children. The trial was registered at clinicaltrials.gov (NCT04774991).

We conducted this analysis from the perspective of the government health sector, specifically the Ministère de la Santé Publique (Ministry of Health) in Niger. We focused on financial costs, although we also do a sensitivity analysis to evaluate elements that carry a non-financial economic burden. We extracted the majority of cost data from the delivery trial expenditure records, with exceptions detailed below. Operational costs likely to represent ongoing program costs were included, and research-specific costs were excluded, based on expenditure records and as necessary consultations with the local team. Specifically, the expenditures that were excluded comprised wages for the post-distribution survey team, per diem for the medical consultant, and the costs of stringent drug monitoring, the latter being necessary for the concurrent mortality trial but unlikely to be implemented in a real-world MDA. We estimated costs for a single calendar year that includes two rounds of drug distribution. We report costs in 2025 USD. As the Delivery I trial was conducted in 2021, we first updated all costs except CHW wages using the inflation rate over that time.^17^ CHW wages were not inflated as they are standardized in Niger and have not changed since 2021. Costs were converted from West African CFA at a commercial exchange rate of 561.88 CFA per USD.^18^

We report cost outcomes at three scales: national, regional, and per dose delivered. The target population for the national program is children aged 1-59 months living in non-urban communities with populations of less than 2,500, following the eligibility criteria of the overall AVENIR project for trial purposes. Population census data from the Niger government for 2021 were used to determine number of children 1-59 months in each region.^19^ The 2021 census data did not include breakdown by community so we used 2012 data^20^ for community number and size as that was the most recent collection of these data. Due to contradicting trends of population growth with depopulation of rural areas since 2012, we could not confidently update the number of eligible communities. For our primary analysis we assumed 90% coverage with two doses annually, based on coverage achieved in the AVENIR Delivery I trial.^10^ The number of Centres de Santé Integré (CSIs, primary health care centers) and districts in Niger was received from the Nigerien District Health Information System (DHIS) using data from 2021.

### Cost Estimation and Categorization

We conducted a micro-costing exercise, which calculated the anticipated annual cost of a national program by summing costs associated with each programmatic activity. In most instances, costs were based on recorded expenditures from the trial for personnel, equipment, training, and supplies. These costs were validated through discussions with the local financial team, review of logged receipts for items, and verification by team members. When records or receipts were not available, the team provided local informed estimation, for instance by going to the market to record the current value of a good.

While the AVENIR Delivery I trial occurred alongside a larger randomized clinical trial, costs were estimated as if the program would be a standalone implementation. Therefore, some central costs that were shared between the Delivery I trial and the larger clinical trial were included at their full rate for this estimation. For instance, the budget team served both programs but is represented as fully dedicated to implementation in this estimation.

We report costs in five activity categories: distribution; training; central administration; drug transport and storage; and monitoring and evaluation. Distribution includes equipment and personnel needed for community entry and delivering the drug to children. Training costs were estimated at the district and CSI levels. Central costs encompass administration of the program at national, regional, and district levels. Drug transport and storage costs encompass delivering the drug to communities, as well as drug quality control. The cost of the drug itself is omitted from the base case, as it was donated from Pfizer and continued donation is planned; as a scenario analysis we include purchase and international shipping. Monitoring and evaluation are activities such as a pregnancy history and sample collection for antimicrobial resistance monitoring to ensure quality and safety.

Personnel and equipment costs related to azithromycin distribution to children (“distribution costs”) were based on expenditures and unit prices observed during the distribution which are detailed in the supplemental material. Distribution costs included two rounds of drug delivery per year. For personnel, we estimated that at least one CHW would be deployed per community, with additional CHW estimated at one per 500 population for communities larger than 500. This reflects operational plans by the Nigerien Ministry of Health. However, CHW are not always available in every community to participate in the MDA. To cover 26% of the CHW needs in AVENIR Delivery I distribution, a CHW was hired from a neighboring district to conduct the MDA. In these instances, a local mobilizer or guide was also hired to facilitate the door-to-door delivery. This extra expense is partially offset by reduced training costs, as the external CHW will have already received training in their own community.

Personnel are compensated via *per diems*. Village and CSI chiefs are given one *per diem* payment per round, and mobiliser/guides, case agents from the CSI, and CHWs receive per diem for two days of working per round (Supplemental Table 1). CHWs are given a transportation per diem once per round regardless of how many villages they visit. CSI chiefs are given a communication per diem once per round. Backpacks and dosing poles are provided once per year. Hand sanitizer is given to every CHW at each round. Mineral water is used to reconstitute the azithromycin powder for oral suspension, and the number of bottles needed was estimated using an average 3.6 doses per bottle of azithromycin and 32 bottles of azithromycin diluted per bottle of mineral water. Syringes are needed for infants under one year of age and calculated as one per infant per round.

Sensitization activities to communicate and educate about the MDA program are also included as distribution costs. A sensitization occurs before every MDA distribution. The first sensitization of the year occurs at the district level, with national, regional, district, CSI, and community level representatives all being compensated for their time and provided with refreshments. A radio program similar public communications also occur as a yearly expense. The second MDA distribution involves a smaller scale sensitization that just includes CSI and community leaders being compensated for their time. Mapping exercises are also conducted annually in order to update information on eligible communities and plan operations, for which participants are compensated for their time. Regional estimates for distribution costs reflect the application of costs for a single child, community, or CSI across their region-specific counts.

Training costs include those related to the entire cascade in a training-of-trainers model, where district and CSI leaders received training one day, and then provided training to the CHWs under their jurisdiction on another day. Each level of training took a single day. The first level of training occurred in each district and included a national supervisor who was given a per diem and a room rental. District chiefs were provided with a communication budget to coordinate the training, as well as a per diem, meals and coffee. A district communicator was also given a budget for communicating to the CSI chiefs about attendance. CSI chiefs were provided with per diem, transportation costs, communication budget, meals, and coffee. On the second day of training, CSI chiefs trained the CHWs, as well as case agents from the CSI. CSI chiefs were provided with a per diem, meals, and coffee. Case agents were given per diem, meals and coffee, as well as a transportation budget. CHWs received the same plus notebooks and pens. As some CHWs will cover more than one community (see paragraph above), we calculated the number of CHWs attending training as 74% of the estimate if all CHW were working in their home community. Training occurs prior to each round, as there can be substantial turnover in healthcare professionals, and also as a refresher for personnel who have previously been engaged in azithromycin distribution.

Central costs were first estimated for each level of health administration (national, regional, and district), then multiplied by the count of administrative units at each level. The national scale up is run by the same non-profit organization, Centre de Recherche et Interventions en Santé Publique (CRISP), that ran the trial so these costs will remain the same as in the trial. These counts were based on 2012 census data (RENALOC) and 2021 DHIS data (Table 1). For costs at the national level, costs were split evenly between all regions. All central costs were calculated by multiplying the observed unit cost by the quantity needed annually. Total counts of geographic information including number of districts, CSIs, population numbers, and numbers of CHWs for each geographic level.

**Table 1.** Geographic information of Niger.

| Region | CSI | Districts | Under 5 population | Non-urban communities | Small grappes < 200 population | CHWs (per-capita scenario) | CHWs (2 relais scenario) |
| --- | --- | --- | --- | --- | --- | --- | --- |
| **Dosso** | 183 | 8 | 557,739 | 4,919 | 2,623 | 4,722 | 7,281 |
| **Tahoua** | 266 | 13 | 905,631 | 4,130 | 1,345 | 4,779 | 6,113 |
| **Maradi** | 209 | 9 | 922,828 | 5,420 | 1,654 | 6,227 | 8,022 |
| **Zinder** | 219 | 11 | 961,425 | 8,414 | 3,929 | 8,126 | 12,453 |
| **Tillaberi** | 278 | 13 | 739,661 | 6,011 | 2,490 | 6,014 | 8,897 |
| **Agadez** | 94 | 7 | 127,247 | 1,216 | 765 | 997 | 1,800 |
| **Diffa** | 72 | 6 | 155,009 | 2,632 | 1,942 | 2,124 | 3,896 |
| **National** | 1,321 | 67 | 4,369,540 | 32,742 | 14,478 | 32,989 | 48,462 |

Drug transportation and storage costs were provided via quotes from the Niger National Office for Pharmaceutical and Chemical Products (ONPPC). The cost of drug reception, inventory, and transportation to every CSI was given as a flat cost specific to each region. The cost of quality control, which includes verification of the volume, quantity, label, and chemical composition of one bottle per lot, for each treatment was estimated as a per-lot cost. We estimated that a national program would require 10 lots per distribution. The cost for drug storage was given as a rate per cubic meter per month, with each cubic meter holding one pallet of drugs containing 3,840 bottles. The number of pallets needed was estimated using the number of doses required per region, assuming each bottle would give 3.6 doses, and rounded up to whole pallets per region. These costs were then summed at the regional and national levels.

The category of monitoring and evaluation includes the cost of surveillance for antimicrobial resistance (AMR) as well as mortality through pregnancy history data collection. In this case, we departed from micro-costing and estimated the fraction of the overall AVENIR expenditures that were dedicated to AMR monitoring, then applied this fraction as a multiplier on the estimated budget for all other activities. AMR monitoring costs comprised 5% of total AVENIR expenditures, so we included it in the budget as 5% of the cost total for all other categories.

Similarly, we included mortality monitoring cost in the same way, as 5% of the total cost for all other categories, not including (or including) AMR monitoring costs. Sensitization activities to educate and communicate with leaders regarding these data collections are also included in mortality and evaluation costs.

We were also interested in “cross-cutting categories” of the fraction of expenditure that would be spent on transportation, education, information, and communication. Transportation costs include CHW transportation for distribution, transportation for all groups (CHWs, case agents, and CSI chiefs) for training, vehicle cost, maintenance, and driver salaries for central costs, as well as cost to bring treatment to CSIs (labeled as drug transport). Education, information, and communication costs include communication costs for CSI chiefs during distribution, district chief and CSI chief communication costs during training, and sensitization costs for AZ-MDA.

### Scenario Analysis

The AVENIR Delivery I trial deployed two CHWs per community for each distribution. With upcoming healthcare system reforms, nationwide implementation may instead assign a number of CHWs based on size of the community. We’ve included a comparison of costs from these two scenarios to see how the decision of how many CHWs to use will impact the costs. The primary scenario assumes one CHW distributing azithromycin per 500 population in each community, rounded up. The second scenario assumes two CHWs distributing azithromycin per community regardless of community size. In both analyses, we specified that 26% of the CHW would come from a neighboring community, as described above. Quantities for equipment specific to an individual CHW are also adjusted as the number of distinct CHW changes. Although the rebalancing of personnel modeled for Scenario 1 may improve coverage compared to Scenario 2, we did not have empirical data to support different coverage between these scenarios.

To address uncertainty regarding the exact procedures for national AZ-MDA implementation in Niger, we analyzed six programmatic scenarios to determine their impact on the overall budget and the percent change in cost. These include: changing the coverage (to 80% and 100%); including drug price at a low-end ($0.40 per dose) and a high-end ($0.70 per dose); removing notebooks and pens for the CHWs during training; reducing the percentage of central administrator salaries paid at the regional and district level to 50% as their time may be shared with other programs; reducing CHW training costs by 50% as the training time may be shortened or consolidated with other programs; and varying the percentage of communities with local CHWs available to work during the distribution.

### Sensitivity Analysis

We conducted a series of one-way sensitivity analyses to identify the budget areas where a change would have the most dramatic budget impact. We separated costs into more volatile and less volatile and tested a 50% increase or decrease compared to baseline for more volatile groups, and a 25% increase or decrease for less volatile groups. More volatile groups included cost of fuel for vehicles, vehicle costs, and drug transport and storage costs; these were deemed more volatile due to occasional shortages in the region.^21^ Less volatile groups included the per diem given to workers involved in training and distribution; equipment needed for distribution; salaries of central administrators; and communication costs used during training.

With our observations of CHW availability and program coverage, we propagated the empirical uncertainty into measures of uncertainty in our estimate. The number of CHWs available in each community varies, which determines the number of mobilizer/guides needed. To determine the bounds for number of mobilizer/guides needed, the 95% confidence interval for proportion of mobilizers was calculated using a Beta distribution with shape parameters α = 20 and β = 58 to simulate uncertainty centered around the 26% mobilizer use seen in AVENIR Delivery I.^10^

Another factor that influences the uncertainty interval is the treatment coverage. Costs that are influenced by treatment coverage include the number of bottles of mineral water and syringes needed for distribution, as well the cost for storage of drug. The cost per dose is also influenced, as the number of doses delivered in the denominator is dependent on coverage. To determine 95% confidence interval bounds for treatment coverage, treatment coverages at the community level seen in AVENIR Delivery I for the door-to-door modality at Round 2 was used, capping treatment coverages at 100%.

## RESULTS

The national budget for one year of biannual azithromycin distribution to treat children aged 1-59 months in Niger is estimated to be $12.50M (95% UI $12.22M – $13.04M) (Table 2).

**Table 2.** Number of doses delivered, overall costs, and cost per dose of the MDA program by category.

| **Cost Type** | **Dosso**<br>**Estimate**<br>**(95% UI)** | **Tahoua**<br>**Estimate**<br>**(95% UI)** | **Maradi**<br>**Estimate**<br>**(95% UI)** | **Zinder**<br>**Estimate**<br>**(95% UI)** | **Tillaberi**<br>**Estimate**<br>**(95% UI)** | **Agadez**<br>**Estimate**<br>**(95% UI)** | **Diffa**<br>**Estimate**<br>**(95% UI)** | **National**<br>**Estimate**<br>**(95% UI)** |
| --- | --- | --- | --- | --- | --- | --- | --- | --- |
| **Number of**<br>**Doses** | 1,003,930<br>(725,061,<br>1,115,478) | 1,630,136<br>(1,177,320,<br>1,811,262) | 1,661,090<br>(1,199,676,<br>1,845,656) | 1,730,565<br>(1,249,853,<br>1,922,850) | 1,331,390<br>(961,559,<br>1,479,322) | 229,045<br>(165,421,<br>254,494) | 279,016<br>(201,512,<br>310,018) | 7,865,172<br>(5,680,402,<br>8,739,080) |
| **Distribution** |  |  |  |  |  |  |  |  |
| **Overall cost** | \$870,334<br>(\$846,357,<br>\$906,395) | \$851,239<br>(\$823,659,<br>\$882,296) | \$1,067,483<br>(\$1,034,426,<br>\$1,106,573) | \$1,462,351<br>(\$1,421,954,<br>\$1,512,042) | \$1,105,443<br>(\$1,074,953,<br>\$1,142,646) | \$224,645<br>(\$218,527,<br>\$231,848) | \$437,113<br>(\$426,588,<br>\$450,631) | \$6,018,609<br>(\$5,846,464,<br>\$6,232,430) |
| **Cost per**<br>**Dose** | \$0.87<br>(\$0.76,<br>\$1.25) | \$0.52<br>(\$0.45,<br>\$0.75) | \$0.64<br>(\$0.56,<br>\$0.92) | \$0.85<br>(\$0.74,<br>\$1.21) | \$0.83<br>(\$0.73,<br>\$1.19) | \$0.98<br>(\$0.86,<br>\$1.40) | \$1.57<br>(\$1.38,<br>\$2.24) | \$0.77<br>(\$0.67, \$1.10) |
| **Training** |  |  |  |  |  |  |  |  |
| **Overall cost** | \$291,651<br>(\$285,487,<br>\$328,631) | \$315,229<br>(\$308,990,<br>\$352,661) | \$373,298<br>(\$365,170,<br>\$422,067) | \$469,546<br>(\$458,939,<br>\$533,189) | \$387,651<br>(\$379,801,<br>\$434,751) | \$82,736<br>(\$81,435,<br>\$90,537) | \$129,536<br>(\$126,763,<br>\$146,171) | \$2,049,647<br>(\$2,006,586,<br>\$2,308,007) |
| **Cost per**<br>**dose** | \$0.29<br>(\$0.26,<br>\$0.45) | \$0.19<br>(\$0.17,<br>\$0.30) | \$0.22<br>(\$0.20,<br>\$0.35) | \$0.27<br>(\$0.24,<br>\$0.43) | \$0.29<br>(\$0.26,<br>\$0.45) | \$0.36<br>(\$0.32,<br>\$0.55) | \$0.46<br>(\$0.41,<br>\$0.73) | \$0.26<br>(\$0.23, \$0.41) |
| **Central** |  |  |  |  |  |  |  |  |
| **Overall cost** | \$362,380<br>(\$362,380,<br>\$362,380) | \$512,794<br>(\$512,794,<br>\$512,794) | \$392,463<br>(\$392,463,<br>\$392,463) | \$452,628<br>(\$452,628,<br>\$452,628) | \$512,794<br>(\$512,794,<br>\$512,794) | \$332,297<br>(\$332,297,<br>\$332,297) | \$302,214<br>(\$302,214,<br>\$302,214) | \$2,867,571<br>(\$2,867,571,<br>\$2,867,571) |
| **Cost per**<br>**dose** | \$0.36<br>(\$0.32,<br>\$0.50) | \$0.31<br>(\$0.28,<br>\$0.44) | \$0.24<br>(\$0.21,<br>\$0.33) | \$0.26<br>(\$0.24,<br>\$0.36) | \$0.39<br>(\$0.35,<br>\$0.53) | \$1.45<br>(\$1.31,<br>\$2.01) | \$1.08<br>(\$0.97,<br>\$1.50) | \$0.36<br>(\$0.33, \$0.50) |
| **Drug**<br>**Transport** |  |  |  |  |  |  |  |  |
| **Overall cost** | \$32,055<br>(\$27,305,<br>\$33,955) | \$57,210<br>(\$49,609,<br>\$60,535) | \$66,027<br>(\$57,951,<br>\$69,115) | \$75,827<br>(\$67,514,<br>\$79,153) | \$84,875<br>(\$78,462,<br>\$87,488) | \$66,760<br>(\$65,573,<br>\$67,236) | \$46,396<br>(\$44,971,<br>\$46,871) | \$429,151<br>(\$391,386,<br>\$444,352) |
| **Cost per**<br>**dose** | \$0.03<br>(\$0.02,<br>\$0.05) | \$0.04<br>(\$0.03,<br>\$0.05) | \$0.04<br>(\$0.03,<br>\$0.06) | \$0.04<br>(\$0.04,<br>\$0.06) | \$0.06<br>(\$0.05,<br>\$0.09) | \$0.29<br>(\$0.26,<br>\$0.41) | \$0.17<br>(\$0.15,<br>\$0.23) | \$0.05<br>(\$0.04, \$0.08) |
| **Monitoring** |  |  |  |  |  |  |  |  |
| **Overall cost** | \$155,642<br>(\$152,153,<br>\$163,136) | \$173,647<br>(\$169,505,<br>\$180,829) | \$189,927<br>(\$185,001,<br>\$199,022) | \$246,035<br>(\$240,104,<br>\$257,701) | \$209,076<br>(\$204,601,<br>\$217,768) | \$70,644<br>(\$69,783,<br>\$72,192) | \$91,526<br>(\$90,054,<br>\$94,589) | \$1,136,498<br>(\$1,111,201,<br>\$1,185,236) |
| **Cost per dose** | \$0.16<br>(\$0.14,<br>\$0.22) | \$0.11<br>(\$0.09,<br>\$0.15) | \$0.11<br>(\$0.10,<br>\$0.17) | \$0.14<br>(\$0.12,<br>\$0.21) | \$0.16<br>(\$0.14,<br>\$0.23) | \$0.31<br>(\$0.27,<br>\$0.44) | \$0.33<br>(\$0.29,<br>\$0.47) | \$0.14<br>(\$0.13, \$0.21) |
| **Overall** |  |  |  |  |  |  |  |  |
| **Overall cost** | \$1,712,062<br>(\$1,673,682,<br>\$1,794,497) | \$1,910,119<br>(\$1,864,557,<br>\$1,989,114) | \$2,089,198<br>(\$2,035,011,<br>\$2,189,239) | \$2,706,389<br>(\$2,641,140,<br>\$2,834,713) | \$2,299,839<br>(\$2,250,612,<br>\$2,395,447) | \$777,083<br>(\$767,615,<br>\$794,110) | \$1,006,786<br>(\$990,590,<br>\$1,040,476) | \$12,501,475<br>(\$12,223,207,<br>\$13,037,596) |
| **Cost per dose** | \$1.71<br>(\$1.50,<br>\$2.47) | \$1.17<br>(\$1.03,<br>\$1.69) | \$1.26<br>(\$1.10,<br>\$1.82) | \$1.56<br>(\$1.37,<br>\$2.27) | \$1.73<br>(\$1.52,<br>\$2.49) | \$3.39<br>(\$3.02,<br>\$4.80) | \$3.61<br>(\$3.20,<br>\$5.16) | \$1.59<br>(\$1.40, \$2.30) |

Regional costs vary from $777K (95% UI $768K– $794K) in Agadez to $2.71M (95% UI $2.64M – $2.83M) in Zinder, which are also the least and most populous regions, respectively.

Nationally, there are an estimated 4,369,540 children eligible for treatment (Table 1). The national average cost per dose delivered at a treatment coverage of 90% is $1.59 (95% UI $1.40 – $2.30) (Table 2). Across regions, the cost per dose ranges from $1.17 (95% UI $1.03 – $1.69; Tahoua) to $3.61 (95% UI $3.20 – $5.16; Diffa), differing due to different population sizes and number of districts and/or CSIs.

### Costs by category

Distribution costs comprise the largest fraction of the budget, both nationally and in every region except Agadez (Table 2). The national distribution cost is estimated to be $6.02M (95% UI $5.85M – $6.23M), which is 48.1% of the total budget. Regional distribution costs vary, from $225K (95% UI $219K – $232K) (Agadez) to $1.46M (95% UI $1.42M – $1.51M) (Zinder).

Distribution costs by item are shown in Supplemental Table 1.

The cost of two rounds of training during one year for azithromycin distribution nationally is estimated to be $2.05M (95% UI $2.01M, $2.31M) (Table 2). Regional costs vary, from $83K (95% UI $81K – $91K) (Agadez) to $470K (95% UI $459K – $533K) (Zinder). Training costs by item are shown in Supplemental Table 2. The total drug transport and cold chain storage cost nationally is estimated to be $429K (95% UI $391K - $444K) with regional costs varying from $32K (95% UI $27K - $34K) (Dosso) to $85K (95% UI $78K - $87K) (Tillaberi) (Table 2). The main driver of cost in this category is the distance of the region from the national capital of Niamey. Supplemental Table 3 shows drug transport and cold chain storage cost by item.

The total central cost nationally is estimated to be $2.87M with regional costs varying from $302K (Diffa) to $513K (Tahoua and Tillaberi) (Table 2). Central costs are largely driven by the number of districts in each region, as district personnel are compensated at numerous points during AZ-MDA. We captured no uncertainty in central costs. Central costs by item are shown in Supplemental Table 4. The total monitoring cost nationally is estimated to be $1.14M (95% UI $1.11M - $1.19M) with regional costs varying from $71K (95% UI $70K - $72K) in Agadez, to $246K (95% UI $240K - $257K) in Zinder. Monitoring & evaluation costs are shown in Supplemental Table 5. Nationally, 17.5% (95% UI 17.3% - 18.6%) of the total budget is spent on transportation, and 14.5% (no variance to create an uncertainty interval) on education, information, and communication (Supplemental Table 6).

### Scenario Analysis

We compared costs when assignment of CHW is based on community population size (Scenario 1) against assignment of two CHW per community regardless of population size (Scenario 2) (Figure 1, Supplemental Figure 1). Nationally, assigning the number of CHWs based on community size has 13.2% lower overall cost compared to Scenario 2, with reduced costs specifically during distribution (19.5% lower than Scenario 2) and training (11.9% lower than Scenario 2). The reduced cost for Scenario 1 arises due to many small communities being assigned only one CHW to deliver the drug, which is not offset by populous eligible communities being assigned additional CHW.

**Figure 1.**
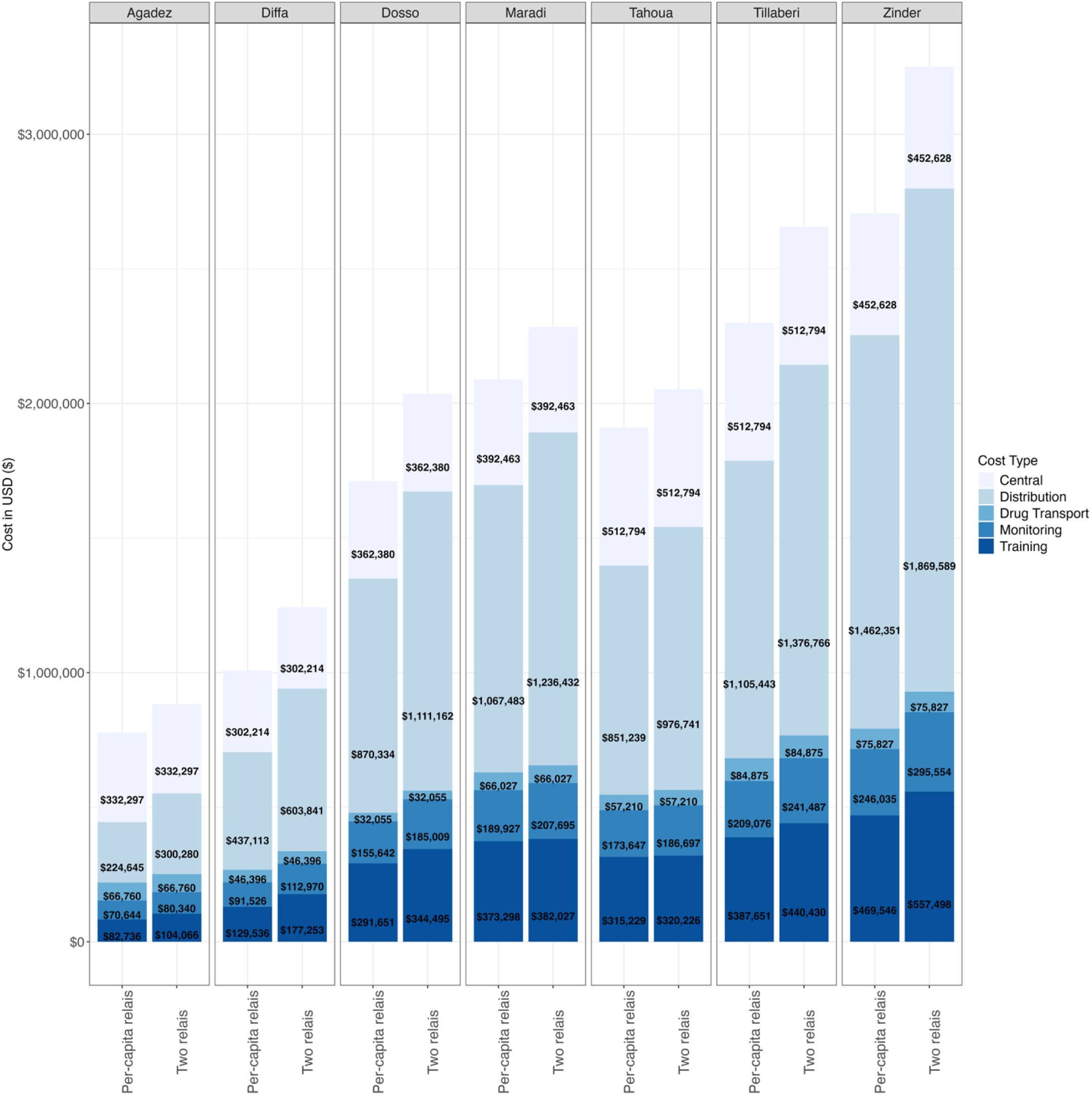
Comparison of costs between two scenarios regarding number of CHWs used in distribution. Differences in shading indicate different cost types. Costs are in 2021 USD.

Six other scenario analyses were conducted (Table 3, Figure 2). First, adjusting treatment coverage to 80% (compared to 90% in the main analysis) decreased the total budget by $36K (0.29%) and increased the cost per dose by $0.19 (12.0%). Adjusted treatment coverage to 100% increased the total budget by 35K (0.28%) and decreased the cost per dose by $0.16 (10.0%).

**Figure 2.**
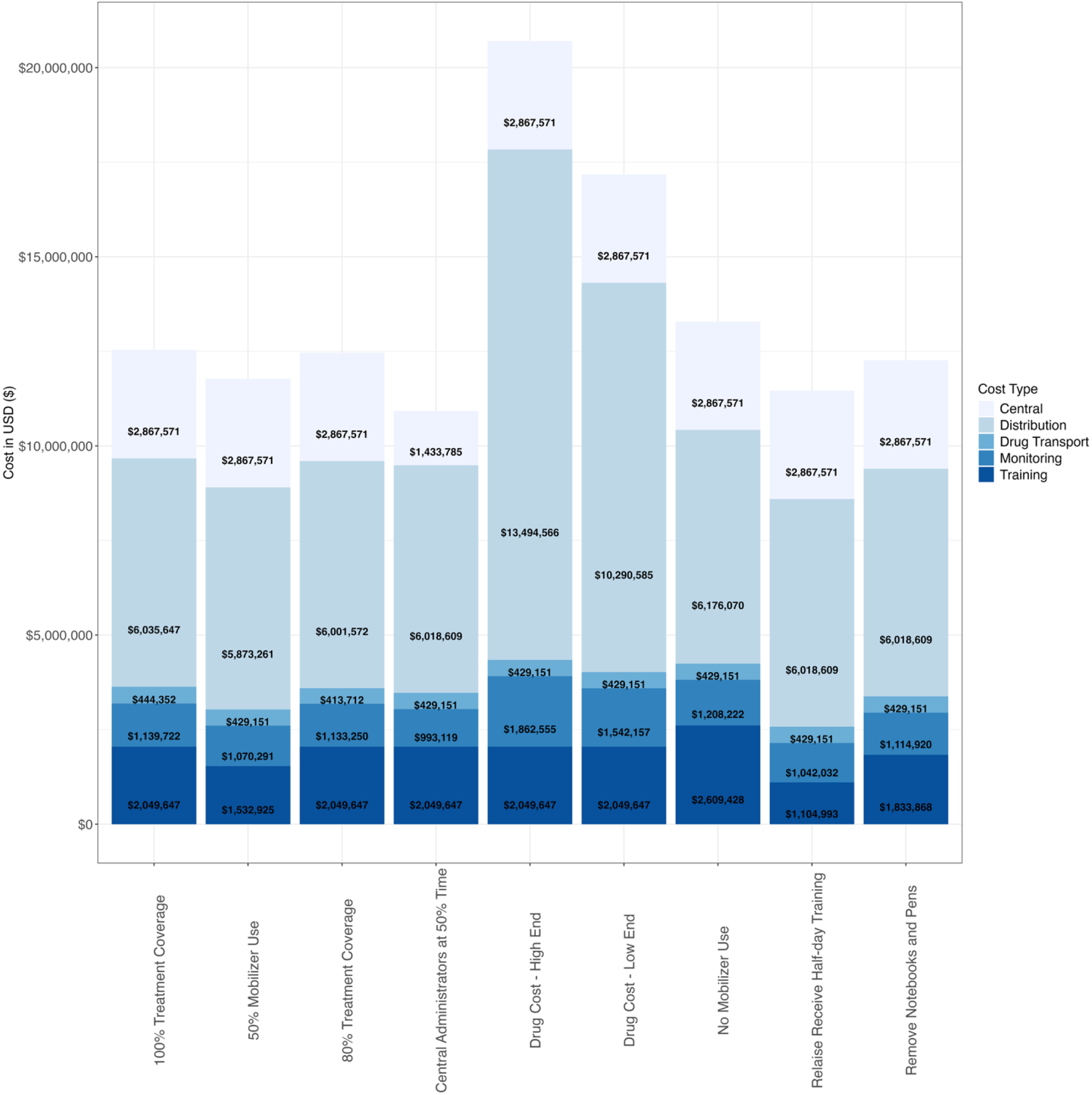
Comparison of costs between scenario analyses. Differences in shading indicate different cost types.

**Table 3.** Scenario analyses including new total budget, cost per dose, and percent changes for each scenario.

| Scenario | New Total Budget |  | New Cost per Dose |  |
| --- | --- | --- | --- | --- |
|  | Budget | Percent Change | Cost per Dose | Percent Change |
| **Change in coverage** |  |  |  |  |
| 80% coverage | \$12,465,751 | -0.3% | \$1.78 | 12.0% |
| 100% coverage | \$12,536,938 | 0.3% | \$1.43 | -10.0% |
| **Add Drug Cost** |  |  |  |  |
| Low-end \$0.40 per dose | \$17,179,110 | 37.4% | \$2.18 | 37.2% |
| High-end \$0.70 per dose | \$20,703,489 | 65.6% | \$2.63 | 65.5% |
| **Relais Receive Half-day Training** | \$11,462,357 | -8.3% | \$1.46 | -8.1% |
| **Remove Notebooks & Pens** | \$12,264,118 | -1.9% | \$1.56 | -1.9% |
| **Central Administrators at 50% Time** | \$10,924,311 | -12.6% | \$1.39 | -12.5% |
| **Change in Mobilizer Use** |  |  |  |  |
| No Mobilizer Use | \$13,290,441 | 6.3% | \$1.69 | 6.3% |
| 50% Mobilizer Use | \$11,773,199 | -5.8% | \$1.50 | -5.6% |

The cost per dose for drug procurement is estimated to be between $0.40 and $0.70 per dose. Including this cost would increase the total budget by $468K (37.4%) to $820K (65.6%). The cost per dose would increase by $0.59 (37.2%) to $1.04 (65.5%) over costs without drug procurement (Table 3).

In our base case reflecting the AVENIR Delivery I trial, CHWs would be given a full day of training dedicated to AZ-MDA. However, CHW training may be able to integrate with other programs in the future. Under a scenario where training for CHWs was limited to only half a day, the national budget decreased by $104K (8.3%) and the cost per dose by $0.13 (8.1%) (Table 3).

In another potential training modification, implementing teams may choose not to provide pens and notebooks to CHW during training, particularly if training for AZ-MDA is consolidated with training for other programs. Removing these notebooks and pens would decrease the national budget by $237K (1.9%) and decrease the cost per dose by $0.03 (1.9%) (Table 3).

In our base case we assumed that there would be central administrators dedicating 100% of their time to this program. However, there is the possibility that these administrators could split their time between this program and others, particularly once the program has been established. To determine the financial impact of this reduced effort, we considered a scenario with regional and district administrators giving 50% of their time to this program, with national administrators remained fully dedicated to AZ-MDA. This change decreased the overall budget by $1.58M (12.6%) and the cost per dose by $0.20 (12.5%) (Table 3).

In our final scenario analysis, we varied the percentage of CHW personnel need that is able to be filled by a local CHW, with the remainder requiring a mobilizer/guide for accompaniment.

Our base case incorporated the observation that 74% of the required personnel needs in AVENIR Delivery I trial were filled by CHWs who reside in the communities they are serving. As AZ-MDA expands beyond the original regions served, this number could change. We varied this percentage from 0 to 100% in intervals of 10% (Table 3, Supplemental Figure 1), finding that the overall cost decreases as CHWs are shared across communities. This reduction is largely driven by need to train fewer CHWs.

### Sensitivity Analysis

To identify the types of expenditures whose cost variation might have the greatest impact on the total budget, we examined changes, both negative and positive, in per diem rates, salaries, equipment, vehicles, fuel, drug transport & storage, and communication (Figure 3). A 25% change in per diem would result in the largest change to the national budget at $846K. Adjustment to fuel charges by 50% would also bring a substantial change at $349K. A 25% change in communication costs would have the smallest budgetary impact at just $9K.

**Figure 3.**
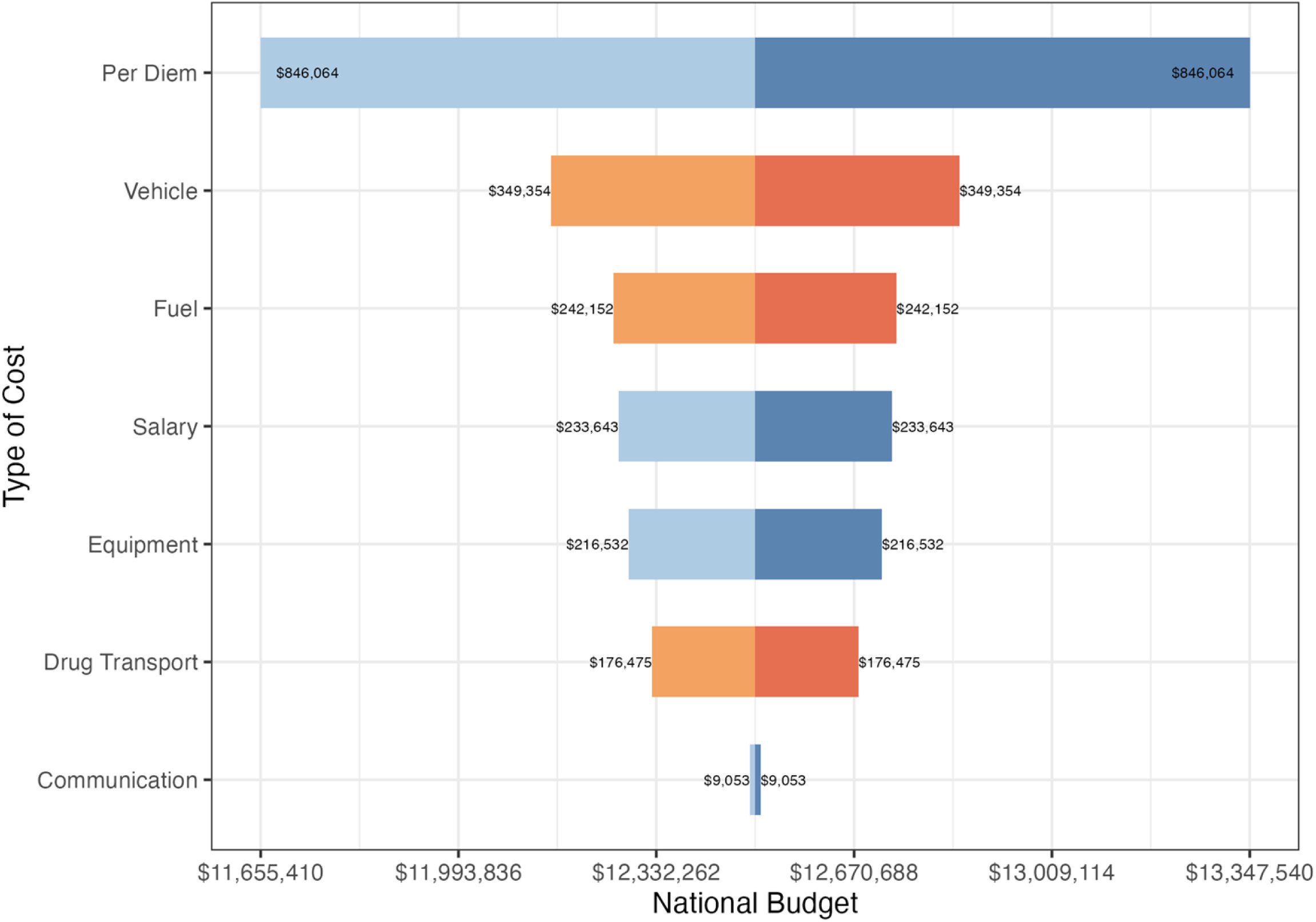
Tornado plot showing the impact of change in selected cost types on the annual national budget for AZ-MDA with low volatility costs with 25% change in blue-scale and high volatility costs with 50% change in orange-scale.

## DISCUSSION

Implementing an AZ-MDA that faithfully replicated the processes used in the AVENIR trials including all rural eligible communities in Niger would cost an estimated $12.50M (95% UI $12.22M - $13.04M) per year, corresponding to $1.59 (95% UI $1.40 - $2.30) cost per dose. We previously published an analysis of training and delivery costs for the AVENIR Delivery I trial, resulting in $1.91 per dose for those categories alone.^10^ However, given the smaller scale of the trial, it is consistent with expectations that it would be less cost-efficient than a national program.

Our cost per dose is higher than found in a previous meta-analysis of MDA against neglected tropical diseases, which identified a benchmark of $0.50 per dose ($0.67 per dose in 2025 dollars).^12^ As noted, AZ-MDA in Niger does not rely on volunteer time, thus raising the cost of the program when compared to the benchmark cost. Furthermore, the meta-analysis identified that cost per dose in rural areas is higher than in urban areas due to the smaller population reached for a similar amount of resources. The AZ-MDA program we model for Niger specifically targets rural communities.

In fact, community size emerges as one of the major drivers of subnational variation in cost per dose for this analysis. When there are many communities with small populations (less than 200 people), we see the cost per dose rise dramatically. For instance, such small communities are over 70% of all eligible communities in Diffa, the most expensive district per dose delivered, compared with less than 1/3 of all eligible communities in Tahoua, the least expensive per dose delivered. This relationship is driven by the fact that each community requires the same activities such as a CHW visit, sensitization, and education – even if there are only a small number of children present as the denominator across which expenses can be spread.

Similarly, the ratio between district supervisory personnel and children is higher when communities are small, driving up central costs. By contrast, children within these small communities may be among those most likely to benefit from AZ-MDA. Small communities are less likely to have their own health center, and there is correlation between distance from a health center and both baseline morality and AZ-MDA effectiveness. Analyses specifically examining the cost-effectiveness of reaching small communities could provide valuable insight for AZ-MDA as well as other rural health programs.

This analysis identifies a few potentially feasible programmatic changes that might increase efficiency without sacrificing treatment coverage. Training for AZ-MDA does not require a full day of work and therefore could potentially be integrated with training for a similar program. Similarly, our assumption that there would be central staff entirely dedicated to AZ-MDA may not reflect reality. If both training and central administration were shared equally with another program, the overall cost could be reduced by up to 20%. However, it may not be easy to identify another program with shared training timing and underutilized central personnel. We recognize as well that this well-funded implementation trial included a high level of supervision, and a long-term program may not choose to sustain this; although this would very likely result in coverage declines. Easier targets such as eliminating branded notebooks and pens do not provide substantial savings and may come with the tradeoff of decreased morale or motivation.

Similarly, our finding that costs are reduced by relying on a smaller cadre of trained CHW accompanied by a larger number of mobilizers might not be an actionable way to achieve savings, if perceived as the deliberate exclusion of trained CHW who reside in a community. Instead, this scenario analysis should be interpreted as a reassurance that there are not negative financial consequences when trained CHWs from neighboring communities are teamed up with mobilizers, as additional distribution expenses are offset by reduced training needs.

A strength of this study is that costs relating to drug distribution and personnel training came from a closely monitored trial. We were able to gather all costs as they occurred, and minimal assumptions were necessary regarding wages, per diems, and market prices. Other costs that weren’t observed within the AVENIR trial such as costs for drug transport and storage came directly from the government of Niger. Therefore, the unit costs included in this budget adhere closely to reality in Niger.

One limitation of our study was uncertainty regarding population size and distribution, as we had to synthesize data sources from different years. Specifically, we calculated the number of CHWs needed in each community based on community population size from 2012 census data, while our costs, number of children under 5 years old, and other geographic information was captured in 2021. There have likely been population changes between 2012 and 2021, however we were not confident making assumptions regarding these changes for rural Niger. Although the national population is growing, it is possible that rural populations are declining due to urbanization trends, so constant growth seemed indefensible. With our costing analytic structure in place, we hope to be able to update the analysis when more recent population data become available.

Across West Africa, trials have been underway to evaluate the impact of azithromycin when delivered in diverse settings and integrated with a variety of platforms.^9,22–25^ The AVENIR trial confirmed azithromycin effectiveness when delivered to children 1-59 months of age, although not when restricted to 1-11 months of age.^9^ Other settings evaluating delivery to only 1-11 months of age have thus far seen no statistically significant effectiveness.^26,27^ Therefore, the expanded age range is likely to be the model for countries considering azithromycin to improve child survival. The MOH in Niger has begun nationwide implementation of AZ-MDA which makes the current analysis extremely salient. As actual expenses from the national program become available, this micro-costing exercise is intended to serve as a source of insight into the drivers of regional differences as well as sources of financial risk. Although other countries will have different population sizes, wage structures, and operational considerations, the costs detailed here can hopefully support informed decisions regarding the deployment of all-to-scarce resources to improve child health and survival in Niger and beyond.

## Data Availability

The datasets analyzed in this study are available in the Open Science Framework (OSF) repository, https://osf.io/hqry6/overview?view_only=98322ff9dc354dc991cebf61a6d36fe2.

## Notes

### Competing Interest Statement

The authors have declared no competing interest.

### Clinical Trial

NCT04774991

### Funding Statement

This work was supported by the Bill & Melinda Gates Foundation grants OPP1210548 and INV-002454.

### Author Declarations

trial approval was obtained from the Institutional Review Boards at the Niger Ministry of Health (Comité Nationale éthique pour la Recherche en Santé) and the University of California, San Francisco.

